# Acceptability, Feasibility, and User Experiences of Continuous Glucose Monitoring in Type 1 Diabetes in Kenya: Perspectives of Adolescents, Young Adults, Caregivers, and Healthcare Providers

**DOI:** 10.64898/2026.08.16.26359665

**Authors:** Agnes Karingo Karume, Prisca Amolo, Lucy Mungai, Hellen Moraa, Titus Arunga, Emmaculate Nzove, Clinton Ndambuki, Lorrein Muhwava, Yvonne Kamau, Elena Marbán-Castro

## Abstract

**Background:** Type 1 diabetes (T1D) is a growing public health concern in low-and middle-income countries, where access to glucose monitoring and consistent routine care remains limited. Continuous glucose monitoring (CGM) may improve diabetes outcomes, but evidence of its acceptability and use in low-resource settings is limited. This study explored the perceptions and experiences of CGM use among people living with T1D, their caregivers, and healthcare providers (HCPs).

**Methods:** Participants were recruited from the ACCEDE-U study, a usability study on CGM use among people living with T1D attending a tertiary referral hospital in Nairobi, Kenya. Among 40 participants in the ACCEDE-U study, those who had completed at least seven weeks of CGM use were eligible to participate in the qualitative component. Three focus group discussions (FGDs) were conducted: one with nine caregivers, one with nine adolescents (12-17 years), and one with six young adults (18-24 years). Semi-structured interviews were conducted with 9 HCPs. Data was collected using guides, audio-recorded, transcribed, and analyzed thematically guided by the socioecological framework. CORE-Q guidelines were used to report results.

**Results:** Participants reported increased engagement in glucose monitoring and high acceptability of CGM. Reduced finger-prick testing and real-time alerts were key benefits, particularly among adolescents and young adults, who valued its discreetness and convenience. CGM was perceived to facilitate sharing of glucose data with HCPs. Caregivers reported a reduced monitoring burden. HCPs perceived CGM as valuable for clinical decision-making by providing real-time insights into glycemic patterns. Cost and limited device availability were identified as major barriers to sustained use.

**Conclusion:** CGM was well accepted and perceived as beneficial. However, challenges related to cost and access may limit broader uptake. Improving affordability and availability could enhance feasibility and promote wider implementation in similar settings.

## Background

Type 1 diabetes mellitus (T1D) is a life-threatening chronic endocrine condition that results in absolute insulin deficiency and requires lifelong insulin therapy^1^. Managing T1D requires constant attention to blood glucose levels, with individuals making frequent daily decisions about insulin use, diet, and physical activity^1^. Without effective diabetes management, people living with T1D are at increased risk of severe complications such as hypoglycemia and diabetic ketoacidosis (DKA), as well as long-term microvascular complications^1^. This ongoing self-management places a substantial burden on patients and their families, particularly in low-and middle-income countries (LMICs) where access to glucose monitoring tools and diabetes care resources may be limited^2,3^. Glucose monitoring enables persons with T1D and their healthcare providers (HCPs) to make informed decisions to optimize glycemic control^4^. For adolescents and young adults particularly, these demands intersect with developmental transitions, increasing independence, and evolving social contexts, often complicating consistent glycemic control^4^.

In resource-limited settings, blood glucose monitoring is usually conducted by self-monitoring of blood glucose (SMBG) using finger-prick testing^5^. However, SMBG does not provide information on blood glucose variability between tests, and self-pricking can be painful and burdensome to people with T1D^6,5^. Continuous glucose monitoring (CGM), which utilizes minimally invasive technology that measures blood glucose every 1-15 minutes, has introduced a shift from SMBG. CGM enables more dynamic glucose monitoring by providing real-time data and glucose trends and patterns over time. It also provides optional alarms for high or low blood glucose readings that alert the person when they are at risk of high or low values^7,8^. Clinical evidence demonstrates that CGM improves glycemic outcomes^2,9,10^ and is now recommended in international guidelines for the management of T1D^11^. The effectiveness of CGM has been shown to be closely linked to how individuals are able to use, interpret, and sustain engagement with the technology in real-world settings^12,13,14^

However, much of the current understanding of CGM use comes from high-income settings, with comparatively little evidence from LMICs^2^. Contextual factors including limited access to devices, cost constraints, and variability in health system support in low-resource settings may influence CGM use^15,16^. To address this evidence gap, larger implementation-research programs have begun evaluating CGM in sub-Saharan Africa, including pragmatic randomized trials of continuous and periodic CGM use in South Africa^17^. Although recent studies from the region have demonstrated that CGM can be implemented and may improve glycemic outcomes, these studies have largely focused on feasibility and clinical endpoints^1^. Emerging qualitative work is beginning to describe how CGM is experienced by users and providers in these settings^18,19^ However, there remains limited qualitative evidence on how CGM is experienced in routine care, particularly among adolescents and young adults with T1D, as well as caregivers and HCPs involved in their care.

Understanding perceptions and experiences is important for identifying barriers to sustained use, as well as factors that support integration of CGM into daily life and clinical care. Whether a health technology is adopted and sustained in routine practice depends not only on its clinical efficacy but also on implementation outcomes such as its acceptability and feasibility for those who use it^28^. Perspectives from users, caregivers, and HCPs are therefore essential in settings where support structures and resource constraints may influence uptake and continued use. This study therefore aimed to explore the acceptability, feasibility, and experiences of CGM among adolescents and young adults with T1D, as well as caregivers and HCPs at the Kenyatta National Hospital (KNH) paediatric and adolescent endocrinology clinic.

## Methods

### Study design and setting

This qualitative study formed part of the mixed-methods usability study (ACCEDE-U) conducted with 40 participants (4-25 years old) living with T1D and attending the paediatric and adolescent endocrinology clinic at KNH in Nairobi, Kenya. KNH is a national referral hospital serving Nairobi County, the wider country, and the region, and is a training institution for the University of Nairobi. The clinics are run by paediatric endocrinologists and diabetes nurse educators.

We used a descriptive qualitative design comprising focus group discussions (FGDs) with people living with T1D and their caregivers, and semi-structured interviews (SSIs) with HCPs. Three FGDs were conducted separately in February and March 2026 with adolescents aged 12-17 years with T1D (n=9), young adults aged 18-24 years with T1D (n=6), and caregivers of children aged 4-12 years with T1D (n=9). SSIs were conducted with HCPs involved in diabetes care and in the review and interpretation of CGM data (n=9), comprising two diabetes nurse educators and seven paediatric medical residents, giving a total of 33 participants.

### Definitions of constructs

The study was framed around four implementation-relevant constructs that structured both data collection and analysis: knowledge and perceptions, acceptability, usability, and feasibility. Acceptability and feasibility were defined in line with the implementation-outcomes taxonomy of Proctor et al^28^. Acceptability was understood as the extent to which participants perceived CGM to be agreeable, satisfactory, and worth using in place of finger-prick self-monitoring. Feasibility was understood as the extent to which CGM could be successfully applied and sustained within participants’ everyday lives, the clinic, and the wider health system, including affordability and availability. Usability was defined, consistent with the parent ACCEDE-U usability study, as the extent to which participants could apply, scan, interpret, and manage the device effectively, efficiently, and with satisfaction. Knowledge and perceptions captured participants’ prior awareness of, beliefs about, and expectations of CGM before and during use. These constructs were examined across the levels of the socioecological framework^20^ (individual, interpersonal, community, and health system/policy) to capture the multilevel factors shaping CGM use.

### Continuous glucose monitoring device

Participants were provided with the FreeStyle Libre 2 system (Abbott Diabetes Care), an intermittently scanned CGM (isCGM) device, for free. Diabetes nurse educators trained participants to apply the sensor themselves. Each sensor was worn for up to 14 days before replacement, during which participants could monitor interstitial glucose continuously. Participants were trained to review their readings regularly, respond to high-and low-glucose alerts, and use the information to guide self-management. At clinic review visits, HCPs downloaded and reviewed the CGM data and glucose trends, discussed the findings with participants, and made insulin adjustments or other care recommendations as needed.

### Recruitment and procedures

Within ACCEDE-U, participants who had been diagnosed with diabetes at least 6 months prior, used CGM continuously for three months and received education on general diabetes care and CGM use prior to sensor application. For this qualitative component, participants were purposively selected to ensure representation of the three key groups (adolescents, young adults, and caregivers). Potential participants were informed about the qualitative study during clinical study visits, and follow-up phone calls were made to confirm availability. Eligible participants were those who had worn the CGM device for at least seven weeks and were available and willing to participate at the time of data collection.

HCPs were eligible if they provided diabetes care at the clinic and reviewed CGM data of at least 10 patients during the study period. HCPs received virtual training on CGM use from the manufacturer and study investigators, followed by hands-on training in interpreting CGM data. Written informed consent was obtained from adult participants, caregivers, and HCPs, while adolescents provided written assent alongside caregiver consent prior to enrolment.

During FGDs, adolescents and young adults described their own experiences using the device, while caregivers shared their perspectives on their children’s use and its impact on diabetes management. During SSIs, HCPs discussed their perceptions of CGM use among children, adolescents, and young adults; their experiences reviewing CGM data and providing careusing CGM information; and their recommendations for implementation.

### Data collection

Discussions took place in a private room within the hospital, with only participants and interviewers present, during participants’ clinic visits. FGDs and SSIs were conducted with guides developed to explore participants’ experiences with CGM, and specifically their knowledge and perceptions, acceptability, usability, feasibility, and recommendations for implementation. The guides were reviewed and refined by the study team prior to data collection.

Two qualitative researchers, HM (female, nurse researcher) and TA (male, social scientist), both trained and experienced in qualitative data collection, facilitated the FGDs; each FGD lasted approximately 45– 60 minutes. SSIs with HCPs were conducted in the clinic at a convenient time and lasted approximately 30 minutes. The interviewers had no prior relationship with participants before study enrolment and were not involved in their routine clinical care. FGDs were conducted in English and Swahili, and HCP interviews in English. All discussions were audio-recorded with consent and transcribed into English. Field notes captured contextual information and key observations.

### Data analysis

We conducted a deductive thematic analysis combining implementation-outcome constructs with the socioecological framework^20^ to understand multilevel factors influencing CGM use. A codebook was developed a priori around the four constructs (knowledge and perceptions, acceptability, usability, and feasibility), and codes were then mapped to the relevant socioecological levels (individual, interpersonal, community, and health system/policy) where they emerged; not all constructs were represented at every level. Two coders independently coded the transcripts. Coding consistency was reviewed iteratively, and discrepancies were resolved through discussion and consensus with a third qualitative researcher. Codes were grouped into themes reflecting facilitators and barriers to CGM use, which were organized in the Results by construct and, within each construct, by the socioecological level(s) at which they emerged from participant narratives. Recurring themes across participant groups suggested that adequate information power had been reached and that the data were sufficient to address the study objectives. All analysis was managed using ATLAS.ti version 24. The study is reported according to the COREQ guidelines^20^.

### Ethical approval

The study was approved by the Kenyatta National Hospital–University of Nairobi Ethics and Research Committee (P291/03/2025).

Additional information regarding the ethical, cultural, and scientific considerations specific to inclusivity in global research is included in the Supporting Information (Inclusivity in Global Research Checklist).

## Results

There were 24 participants included. Nine adolescents (five males and four females) participated in an FGD, with a median age of 14 years (interquartile range (IQR): 13-16). All were students. Nearly half of the adolescents (4/9, 44%) had lived with T1D for five to more than ten years (Table 1).

**Table 1:**
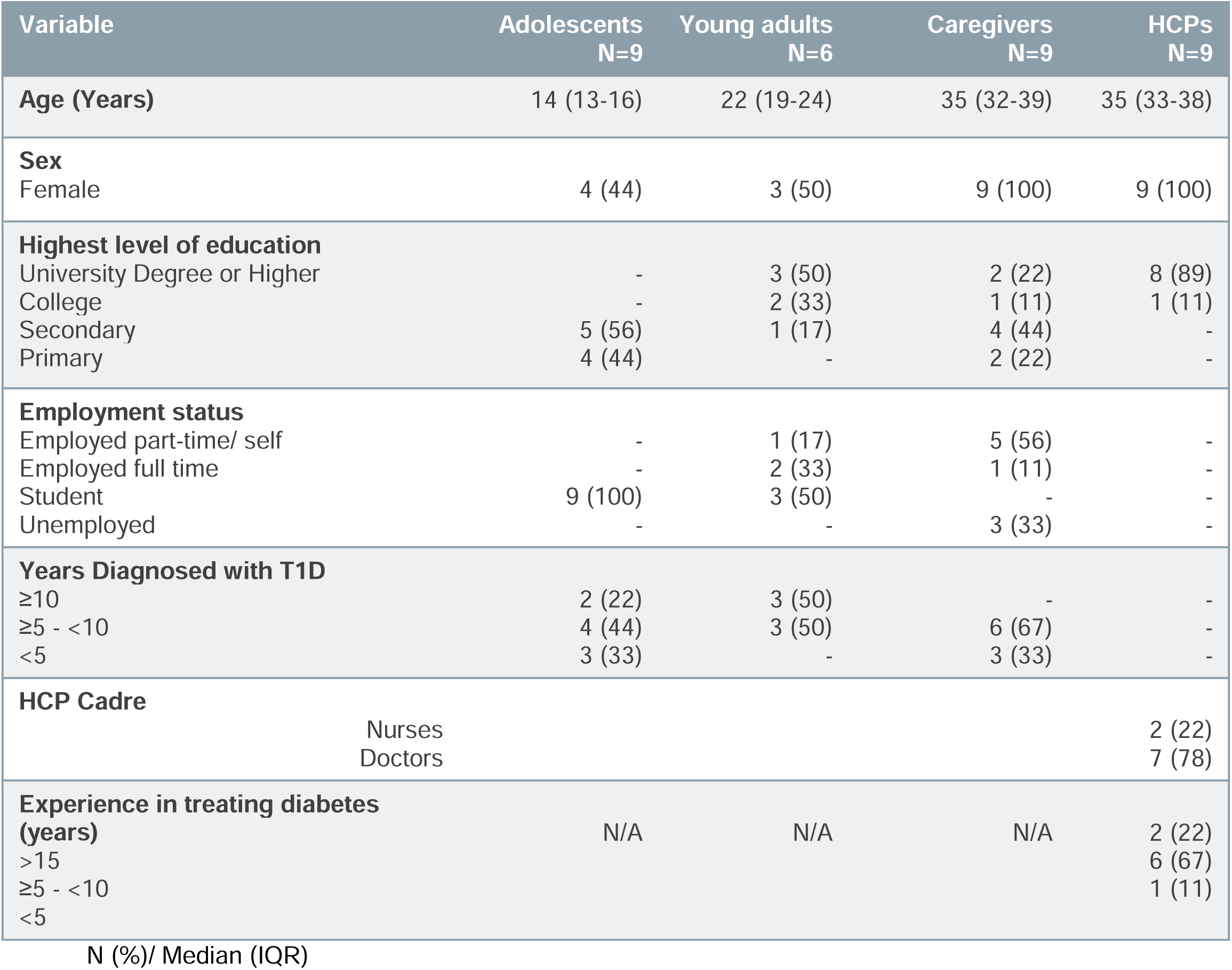
Descriptive characteristics of study participants (N= 33)

Six young adults participated in another FGD, with a median age of 22 years (IQR: 19-24). Three were female and three were male. Half of the young adults had lived with T1D for five to more than ten years, while the other half had T1D for more than 10 years.

A total of nine caregivers, all females, participated in the third FGD, with a median age of 35 years (IQR: 32-39). Most caregivers (6/9, 67%) reported that their children had lived with T1D for five to more than ten years.

A total of nine HCPs participated in the SSIs, including two diabetes nurse educators and seven paediatric medical residents. The median age was 35 years (IQR: 33-38). Most HCPs (6, 67%) had experience treating diabetes for five to more than ten years, while two had over 15 years of experience.

### Knowledge and perceptions of diabetes and continuous glucose monitoring Individual level: Living with TID and personal perceptions of CGM

Caregivers, adults, and adolescents acknowledged that living with T1D was a challenging journey. The challenges described spanned the clinical continuum, beginning with the distress of initial diagnosis and extending to the rigorous demands of daily management, such as dietary modifications, frequent glucose monitoring, and insulin administration. Notably, participants highlighted the profound *“treatment fatigue“* associated with this daily regimen. Some adolescent participants reported periods of therapeutic non-adherence or *“treatment breaks”,* often resuming management only upon health deterioration or external intervention from clinical teams and guardians.

> *“Like for me, it has really affected me in school. When it comes to diet and meals, you eat different foods from the rest, so it makes you feel discriminated against in a way.”* [male adolescent]

Prior to the study, most participants had limited knowledge of CGM. Many had only encountered CGM through social media and believed they were technologies reserved for “*foreigners*” or wealthy families. They admired the device from afar, especially its ability to show blood glucose in trends and allow remote monitoring through a phone but perceived cost as a major barrier to access.

> *“People were saying that it’s expensive, so I knew this thing was for only foreigners. So the thought of a CGM faded.”* [female young adult]

### Interpersonal level: Family support and shared diabetes management

Diabetes management was described as a shared responsibility involving caregivers, family members, schools, and healthcare providers. Caregivers of younger children reported extensive involvement in daily monitoring and insulin administration, often coordinating with teachers and making frequent trips to school to support diabetes care.

> *“So, sometimes it forces some of us to quit our jobs…at times a parent or caregiver is forced to run to school to check their blood sugar level, especially the young children, who cannot inject themselves or even give themselves insulin…”* [female caregiver]

### Community level: Social norms and perceptions surrounding diabetes technologies

Participants’ perceptions of CGM were strongly influenced by wider community narratives and media exposure. Many associated CGM with wealthy individuals, people living abroad or private healthcare.

> *“I used to see them on social media with children abroad and those who are rich. I used to admire it every day because I could see it in that video how they are reading the results. I thought it was only for the rich or those who have some connection with people who are abroad and are able to send them those devices here in Kenya.”* [female caregiver]

### Health system (organizational) level: Healthcare provider knowledge and readiness

Most HCPs reported not having interacted with CGM before the study, and the few with experience cited that they were available in a few private hospital settings. Others had only heard about CGM being available in high-income countries; they assumed it was a device reserved for wealthy patients and not feasible in their setting. Most HCPs reported that they believed CGM was difficult to use, expensive, and complicated to interpret. Others were hopeful that the CGM devices would make it easier both for them and their patients in managing T1D.

> *“I thought the continuous part would be better to evaluate for both us practitioners and the patients, just to see the glucose. Yeah, better than just doing four times a day. That was my assumption, but I didn’t know anything about it.”* [Paediatric resident]

### Acceptability of CGM

#### Indivihdual level: Personal acceptability and perceived benefits of CGM

CGM was widely accepted by caregivers, adolescents and young adults, largely because it removed the struggle and pain associated with repeated finger-pricks. Caregivers described feeling relieved and grateful, noting that children were happier and more willing to engage with their diabetes care than when using glucometers. The overall impression was that the device was easy to use, even for those with limited literacy.

> *“My child fears finger pricking, and we used to quarrel a lot with her. Sometimes, especially during the cold, she could prick, but no blood came out, and also the pain she is feeling. When I heard the news, they were here, and I thanked God. I was so happy. The child also celebrated when she was given the CGM. She was more than happy.”* [female caregiver]
>
> *“First thing, it’s pain-free. I used to hate pricking my fingers. So now it’s pain-free because when you have the sensor there, you just scan.”* [female young adult]

While glucometers were characterized by their logistical burden requiring the coordination of lancets, test strips, and disposal, CGM emerged as a highly acceptable alternative due to its unobtrusive nature. Participants highlighted that CGM eliminated the “*process*” of monitoring, transforming it from a series of disruptive, painful interruptions into a seamless background activity. This reduction in management friction significantly lowered the threshold for engagement, fostering a sense of autonomy and psychological relief compared to the invasive requirements of conventional meters and supplies needed to monitor glucose

> *“The glucometer is time-consuming, and also, when someone falls, the strips will spread all over the place, causing contamination. Again, there could be poor storage of the prickers, and the child might forget to dispose of them well, and they could again cause injury in the house when someone stepped on them or sat where they were placed…glucometer sometimes brings an error…caused by a low battery, and yet to find a replaceable battery is not easy in rural area.”* [female caregiver]

CGM was seen as beneficial in many aspects, such as providing alerts whenever a patient is going into hypoglycemia, which is a state that most are often worried about. Even those around them were impressed by the CGM device’s ability to provide an alert before the hypoglycemia occurs.

> *“I’ve found it convenient and easy to use. It’s okay. For example, when I’m in school, I get hypoglycemia. Before I had a CGM, it used to scare me a lot because it might be low, like 2 or 4, and you’re shaking, and maybe there’s no one else who knows how to read the sugar levels. Still, with this, even my class teacher advised me that its good since if I’m somewhere and even my blood sugar starts to drop, I’ll hear a reminder and look at the alarm and see the levels are down then I’ll get to know what to do or if its high I have to do this, so to me it’s okay.”* [female adolescent]
>
> *“It has helped in sugar level management because of the graphs that track the level of sugar and allow you to take immediate action to stabilize the levels when high or low.”* [male adolescent]
>
> *“I am happy with this CGM, especially at night, because I do not have any more stress when I am sleeping. When the child is going to hypoglycemia, I would not have a problem knowing that the alarm will call when it is heading to hypoglycemia.”* [female caregiver]

#### Interpersonal level: Strengthening caregiver support and shared diabetes management

CGM strengthened interactions between children, caregivers and HCPs by facilitating shared decision-making and improving communication around diabetes management. The ability to scan and immediately obtain glucose readings enabled caregivers to identify missed insulin doses or unreported food intake without relying solely on children’s self-reports.

> *“…when she lies to you, you will just scan and get the result, and from there, I will know where she has come from, even if she has eaten something and failed to inject insulin, I will know.”* [female caregiver]

#### Community level: Community perceptions influencing acceptability

Although CGM was generally well accepted by users, community reactions initially influenced its acceptability. Caregivers reported that the introduction of CGM initially raised suspicion and misunderstanding within their communities. Before the devices were applied, some family members feared that unfamiliar technology could cause harm.

> *“We went and shared with the spouse and other family members, and they were like, ‘Weuh! If I get you with those foreign things, they can bring a problem to the child.’”* [female caregiver]

Some adolescents described feeling self-conscious when wearing CGM in public because of the repeated questions from peers and community members:

> *“There was a time when I wore a short-sleeved shirt and the CGM was visible, so everyone kept on asking about it until I was embarrassed (…). I tried to avoid them and was not responding as much as they kept asking me. You find small kids want to play with it, so I told them it’s not something to play with. Until now, I started lying that when you play with it, it can shock you.”* [male adolescent]

#### Health system (organizational) level: Acceptability among healthcare providers

HCPs reported that their experience with CGM exceeded initial expectations, demonstrating high acceptability within the clinical setting. The continuous glucose data was perceived as a valuable tool for enhancing clinical decision-making by providing detailed, real-time insights into glycemic patterns. Additionally, HCPs noted that access to continuous data supported both patients and caregivers in better understanding and managing glucose levels, further reinforcing the utility of CGM in routine care.

> *“I’m able to give my diabetes education in a better manner, because then I have the real data. Yeah. You know, like the 24-hour thing. Then I’m able, to know what to teach and where to teach. Then I’m able to probe as well. Why are we having this here? What do you think would have happened here? Then we’re able to come up with a solution, together with the patient and the guardian or the parent, for us to have better glucose readings.”* [Diabetes Nurse]
>
> *“Someone can see the sugar levels, time in range when it is very high, the hyperglycemia or hypoglycemia, and also it brings a red indicator when someone is hypoglycemic, indicating that this patient is hypoglycemic at that time. I have been managing some of the patients and this helps me make a clinical judgment based on that.”* [Paediatric resident]

HCPs noticed that some of their patients who previously struggled with glucose control improved significantly. This group was likely motivated by being able to see their ‘real-time’ data at home and being able to discuss the same with their HCPs.

> *“With CGM, we are able to control their blood sugar better…and a better HbA1c is overall a good thing for our patients… it is a good measure to help us avoid many complications that come with diabetes. It would also reduce the number of hospital visits. When the child is having well-controlled sugar in school, they can be alert in school, concentrate, and even perform better.”* [Paediatric resident]
>
> *“When we go to the statistics in our files, I’ve seen the HbA1c improve. Patients who never had an HbA1c below 10, we’ve seen several below 10 because this time they were on CGM, they are able now to see, they are able now to do the scan, it’s motivating, and they are able now to manage themselves better, and so the outcome, it’s good and good, and it can be better and better.”* [Diabetes Nurse]

### Usability of CGM

#### Individual level: Ease of use and user experiences with CGM

Most caregivers, adolescents and young adults reported that CGM was easy to learn and became increasingly intuitive with repeated use. Although participants initially experienced some anxiety when applying the device independently at home, confidence improved over time. Users appreciated the simplicity of scanning the sensor to obtain glucose readings and expressed high satisfaction with the CGM, citing its intuitive interface.

> *“Yes, this CGM is helping a lot because after the child eats something, around 30 minutes or an hour, I should check to know how the child’s sugar level is. This has been so easy to determine for me using the CGM than before when I was using the glucometer. We could just guess or assume because we did not know anything about that. Currently, even the child is happy and wants to know her sugar level after eating by just scanning, and by that, she will be able to know how she is doing.”* [female caregiver]

Participants also appreciated the convenience of CGM particularly during sleep, since glucose levels could be checked without disturbing the user.

> *“CGM has made it very easy. Also, it can notify someone around you about your sugar levels, unlike the glucometer, where someone would first have to wake you up at 3 am to test your sugar. But with CGM, one just comes, lifts you, does a scan to check the sugar levels, and takes the readings. So it was in a simple manner, and it’s user-friendly for everyone to use. I love it so much.”* [male young adult]
>
> *“The good thing with the sensor is that as long as someone knows where the sensor is, they can scan even over the blanket, and it will detect. There is no disturbance, as with the case of the glucometer, whereby the child must be woken up, or when finger pricked, the pain will make him wake up.”* [female caregiver]

Despite these positive experiences, participants reported a few usability challenges. Some reported CGM application site itchiness, change of skin color, and dryness. However, while this was concerning, it did not deter from continued CGM use; they rotated sites in the subsequent CGM application, and often these side effects subsided within a short period.

> *“But I feel like maybe the sensor, putting it on a few patients, the parents complained. It was a bit irritating and hurtful, and it left marks. So I felt like some were not happy with that.”* [Paediatric resident]
>
> *“The tapes react with some people’s skin, for example, they cause rashes like pimples.”* [female caregiver]

Some participants reported occasional sensor failures, loss of signal and premature detachment. When this occurred, faulty sensors were replaced, allowing participants to continue using the CGM.

> *“…Sensor has an error, yet it hasn’t been touched or affected in any way, then the battery durability is not okay, it doesn’t last that long, and during blackouts or when power is lost, you cannot really rely on it.”* [male adolescent]
>
> *“I also experienced the falling off of the sensor after seven days. It was in my second CGM sensor when schools opened. It was telling us that there was no signal, but when you look at the CGM, it was intact. It was not removed, and nothing was wrong; it was completely intact still, and we were wondering what the problem could be”* [female caregiver]

#### Interpersonal level: CGM supporting shared diabetes management

Participants described CGM as strengthening communication and collaboration between patients, caregivers and HCPs. Continuous glucose trends facilitated more meaningful clinical discussions, allowing HCPs and families to jointly identify factors contributing to glucose fluctuations and develop individualized management strategies.

> *“It helps because like for example when I come here to the clinic and they observe the trends of my blood sugars, let’s say its high, they would ask me what I did for it to rise because there are graphs that shows certain points where the graphs are usually high like maybe from 10 −12 and what do I normally do on that particular time.”* [female adolescent]

#### Health system (organizational) level: Healthcare provider experiences supporting CGM usability

HCPs corroborated reports from caregivers, adolescents, and adult participants, noting that initial use of CGM was sometimes accompanied by anxiety and uncertainty, particularly around applying and managing the device at home. HCPs explained that users were often initially worried about whether they would be able to use the device correctly or manage it independently. However, this apprehension reduced over time as users became more familiar with the device and gained confidence through HCP guidance, training, and repeated use.

> *“Initially, it was not easy, again, because I think, because of the anxiety, we do have fallouts and like, am I sure that, you know, like, now that we are going home, will I be able to fix it for my baby? But with time, I mean, as I think towards the end, it was an easier process than it was initially. Again, remember, this is something that is new. Yeah. So with time, it has become easier.”* [Diabetes nurse]

HCPs also reported that CGM results were easy to interpret with minimal training, making CGM an ideal tool for busy clinical teams. Those who had a chance to receive an online training session and got additional material felt it helped them understand the process of CGM application, and how to interpret results.

> *“We were trained, and the session really helped us to know what to do once we interacted with the printout of the glucose levels. The videos, which they also shared with us at some point, and some materials for us to go through before interacting with these patients for the first time.”* [Paediatric resident]

### Feasibility of implementing CGM

#### Individual level: Perceived affordability and sustained use

Participant perceived CGM as highly beneficial for diabetes management but questioned whether continued use would be financially feasible outside the study. Caregivers described the emotional burden of repeated finger-prick monitoring and expressed a desire to continue using CGM if it were affordable. Participants consistently noted that the recurring cost of replacing sensors would make long term use difficult for many families.

> *“…she could prick her fingers and press them hard, but no blood came out. Her fingers are full of injection marks. She has pricked it severely, and sometimes I just sit and wonder if I could be having the money, the child cannot pass through that traumatic experience of injecting herself now and then.”* [ female caregiver]

This was emphasized even by those who had their first CGM experience in the study as a likely barrier to long-term use.

> *“What might make me not continue using the CGM is its cost. It is very expensive, and within fourteen days, someone is supposed to use around fourteen thousand shillings. Meaning in a month, someone will have to use like thirty thousand shillings. With the current situation, someone cannot afford it because could be the person has no job. Again, the salary cannot even be those thirty thousand shillings. So, in short, the CGM is very expensive, and that is what might make many people not continue using it.”* [female caregiver]

#### Interpersonal level: Family support for sustained CGM use

Participants, particularly adolescents, emphasized the importance of family support in sustaining CGM use and integrating diabetes management into daily life. Parents and other family members provided encouragement, assisted with glucose monitoring, reminded users about medication and diet, and helped integrate diabetes management into everyday routines.

> *“My dad has the same problem (Diabetes), and he encourages me. Then, my mom is always there and helps me in measuring my sugar levels, taking my medicine. If I’m in school, she would call to let me know. Then also on the diet, she tells me what to eat.”* [female adolescent]

#### Community levehl: Social support promoting continued use

Participants highlighted the contribution of peers and wider social networks in supporting ongoing CGM use. Friends and family members who understood diabetes provided practical advice when challenges such as sensor detachment, skin irritation or connectivity problems arose. Educating friends and community also created supportive environments that facilitated continued device use.

> *“I have a very supportive mother and very supportive friends. My mother called all my friends at home, cooked food, and told them about everything. So, if I go to my friends’ house, they know.”* [male young adult]

#### Health system and policy level: Enablers of sustained CGM implementation

HCPs considered affordability to be the most important determinant of wider CGM implementation. They noted that the recurring cost of replacing sensors every two weeks would limit routine use for most families unless financing mechanisms or government subsidies were introduced.

> *“I think for me, the main hurdle is just around the price because it is not cheap. I hear the cheapest one is between eight thousand and eighteen thousand Kenyan shillings. Remember, they are changed every two weeks, meaning twice monthly. So that would be the biggest hurdle because in our settings, our patients cannot afford that eight thousand or eighteen thousand after every two weeks.”* [Paediatric resident]

HCPs also emphasized the importance of provider training for successful implementation. Training on CGM use and interpretation of glucose data enhanced HCPs understanding and confidence in using the technology in routine care. They particularly valued the combination of online training and hands-on experience with patients, which facilitated practical application of knowledge. HCPs noted that continued and structured training would further strengthen confidence and support integration of CGM into clinical workflows, thereby enhancing feasibility.

> *“The online and physical training helped us to have first-hand experience with our patients. We can get online training, but without interacting with the actual clients, you cannot understand it.”* [Paediatric resident]

HCPs also highlighted the importance of patient education using simple, accessible educational materials to support understanding and uptake of CGM

> *“Audiovisual materials would be the best because some people do not know how to read. Alternatively, they can make some videos for the patients who are on CGM and share them with them. They will get to watch the video whenever they need some clarification.”* [Paediatric resident]

Participants further suggested that broader implementation would require supportive health policies to improve access to CGM for children living with T1D. They also emphasized the need to strengthen support systems within schools by educating teachers, nutrition staff and other school personnel to support children using CGM.

> *“Some changes in the health policies may be to say that… all children with type 1 diabetes mellitus require continuous glucose monitoring… these children need a lot of support at home and in school… we need to educate our teachers… nutritionists, cooks at school… to be able to support our children… as they continue to use this device.”* [Paediatric Resident]

## Discussion

This study provides contextual evidence on the acceptability, usability and feasibility of CGM among adolescents, young adults, caregivers, and HCPs in a resource limited setting. Living with T1D was described as physically, emotionally, socially and financially demanding with substantial treatment burden associated with repeated finger-prick monitoring, insulin administration and dietary restrictions. Similar challenges have been reported in other studies, where adolescents and caregivers describe T1D management as burdensome and emotionally demanding, often contributing to diabetes distress and treatment fatigue^21, 22^.

Within this context, CGM may help reduce some of the day-to-day burden of diabetes management by minimizing painful finger-prick testing, providing real-time glucose information and supporting more timely self-management. Our findings reinforce growing evidence demonstrating that CGM can reduce diabetes management burden and improve user engagement, particularly among adolescents and young adults with T1D^21, 23^. This study adds to the limited body of African evidence on CGM implementation, alongside recent qualitative work from South Africa^19^.

Adolescence is recognized as a challenging developmental stage for diabetes self-management due to competing social, behavioral and emotional demands, often resulting in suboptimal adherence and poorer glycemic outcomes^21,23^. In our study, adolescents described challenges maintaining frequent glucose monitoring and diabetes routines, reflecting the everyday demands of living with T1D. CGM appeared to ease this burden by replacing repeated painful interruptions with a more seamless and less intrusive process that was easier to incorporate into daily life. Similar findings have been reported in qualitative studies from other settings where adolescents describe CGM as reducing the physical and emotional burden of diabetes management while promoting greater autonomy and self-efficacy^22,23^. Adolescents and young adults also valued the discreetness and convenience of CGM, which may be particularly important in school and social environments where diabetes self-management may sometimes feel visible or stigmatizing. Previous studies similarly suggested that discreet monitoring technologies may improve comfort, adherence, and social confidence among adolescents and young adults with T1D^23^.

Real-time glucose visibility and alerts may also support improved confidence in diabetes management. Prior studies showed that fear of hypoglycemia is common and may negatively affect quality of life and glycemic management^23^. CGM alerts appeared to provide reassurance to adolescents, young adults and caregivers particularly during school hours and nighttime when glucose monitoring is often difficult. Similar observations have been documented in studies where CGM improved confidence in diabetes management and reduced fear associated with nocturnal hypoglycemia^23^. Caregivers additionally reported that CGM enabled closer supervision of insulin adherence and dietary behaviors while reducing the need for disruptive overnight glucose checks, findings that align with studies demonstrating reduced caregiver stress and improved perceived safety with CGM use^24,25^.

Family and social support may also play an important role in facilitating successful CGM use among adolescents and young adults with T1D. Adolescents often rely on caregivers, peers, and wider social networks for emotional reassurance, practical problem solving and support with daily diabetes management, particularly when navigating new technologies. Family engagements and peer support are important facilitators of diabetes self-management and diabetes technology uptake among adolescents with T1D, contributing to improved confidence, adherence and sustained use. In low resource settings where access to formal diabetes support systems may be limited, strengthening family and community support networks may be particularly important for sustained CGM use^24,25^. Although CGM enhanced caregiver oversight of diabetes management, continuous access to glucose data may also alter parent-adolescent relationships. Increased monitoring could reduce adolescents’ sense of autonomy or contribute to conflict if CGM data are used primarily to detect non-adherence.

Family centered education should therefore emphasize collaborative interpretation of CGM data and supportive communication.

Our findings suggest that CGM may have important clinical value in supporting more individualized diabetes care in routine practice. Access to continuous glucose trends may facilitate more targeted diabetes education, earlier identification of glycemic patterns, and collaborative problem solving between HCPs, adolescents and caregivers. Similar benefits have been reported in prior studies, where CGM use enhanced patient-provider communication and supported more personalized diabetes management^22,23,26^. Additionally, evidence from randomized trials and observational studies demonstrate that CGM is associated with improved glycemic outcomes including lower HbA1c and increased time in range among persons living with T1D^2,27^. However, successful integration of CGM into routine care in low resource settings will likely require HCP training, simple patient education approaches, and health system support to strengthen interpretation and use of CGM data in routine workflows.

Despite overall positive experiences, usability challenges remained important considerations for sustained CGM use. Technical issues such as sensor detachment, device malfunction, and minor skin irritations may affect confidence and continued use, particularly during early adoption. Similar concerns related to device wear, adhesive problems, skin reactions, and technical interruptions have been reported in other studies, although these barriers are generally outweighed by perceived benefits^22,23^. These findings highlighted the importance of practical training, troubleshooting support, and clear patient education to facilitate sustained CGM use in routine patient care.

Cost could be a major challenge to the sustained use and broader implementation of CGM in low-resource settings. The recurring cost of sensors and limited insurance coverage may place routine CGM use beyond the reach of many families, raising important concerns about equitable access to diabetes technologies. Similar disparities in access to CGM and other diabetes technologies have been reported globally, particularly in LMICs where affordability, limited supply chains, and inadequate health financing continue to constrain uptake^2^. These findings highlight the need for implementation approaches that extend beyond device provision alone and include supportive financing mechanisms, health policies, and strategies to improve sustained and equitable access.

Broader system-level support may also be important for successful implementation. Studies emphasized the importance of school-based support, caregiver engagement, HCP training and patient education in optimizing diabetes self-management among adolescents with T1D^21,24^.. In settings such as Kenya, strengthening community awareness and integrating diabetes education within schools and routine care systems may help improve long term uptake and sustained use of CGM.

This study has several strengths, including diverse perspectives from adolescents, young adults, caregivers of children with T1D and HCPs, and the ability to situate findings within the context of a usability and feasibility study. Limitations include social desirability bias, and the fact that participants were drawn from a research study, which may not fully mirror routine practice. Also, there was a relatively small number of participants included in the study. Future work should evaluate long term sustainability, implementation outcomes, and cost effectiveness of CGM integration within routine paediatric and adolescent diabetes care in similar settings.

## Conclusions

Overall, our findings suggest that CGM is a feasible and acceptable intervention to supporting T1D management among adolescents, young adults and caregivers in Kenya, with potential benefits for self-management, caregiver reassurance, and clinical care. However, sustained implementation will likely depend on addressing affordability and strengthening the broader systems needed to support routine use, including HCP training, patient education and supportive health and school-based policies. Efforts to improve equitable access to diabetes technologies may be particularly important in low resource settings, where the burden of T1D management remains high.

## Data Availability

The qualitative data generated and analyzed during this study are not publicly available because they contain potentially identifiable participant information. De-identified data are available upon reasonable request.

## Author Contributions

Study conception and design: EM-C, YK, PA and LM1.

Training: PA, LM1, AKK, YK, EM-C and EN

Data acquisition: HM and TA

Coding and analysis: HM, TA and AKK

Supervision and validation: EM-C, YK, AKK, PA and LM1.

Writing original draft: AKK

Review & editing: PA, LM1, HM, TA, EM-C, YK, LM2, EN, CN

